# Extracting Pulmonary Embolism Diagnoses from Radiology Impressions Using GPT-4o: A Large Language Model Evaluation Study

**DOI:** 10.1101/2024.10.14.24315482

**Authors:** Mohammed Mahyoub, Kacie Dougherty, Ajit Shukla

## Abstract

**Background:** Pulmonary embolism (PE) is a critical condition requiring rapid diagnosis to reduce mortality. Extracting PE diagnoses from radiology reports manually is time-consuming, highlighting the need for automated solutions. Advances in natural language processing (NLP), especially transformer models like GPT-4o, offer promising tools to improve diagnostic accuracy and workflow efficiency in clinical settings.

**Objective:** To develop an automatic extraction system using GPT-4o to extract PE diagnoses from radiology report impressions, enhancing clinical decision-making and workflow efficiency.

**Methods:** Two approaches were developed and evaluated: a fine-tuned Clinical Longformer as a baseline model and a GPT-4o-based extractor. Clinical Longformer, an encoder-only model, was chosen for its robustness in text classification tasks, particularly on smaller scales. GPT-4o, a decoder-only instruction-following LLM, was selected for its advanced language understanding capabilities. The study aimed to evaluate GPT-4o’s ability to perform text classification compared to the baseline Clinical Longformer. The Clinical Longformer was trained on a dataset of 1,000 radiology report impressions and validated on a separate set of 200 samples, while the GPT-4o extractor was validated using the same 200-sample set. Post-deployment performance was further assessed on an additional 200 operational records to evaluate model efficacy in a real-world setting.

**Results:** GPT-4o outperformed the Clinical Longformer in two of the metrics, achieving a sensitivity of 1.0 (95% CI: [1.0, 1.0]; Wilcoxon test, *p* < 0.001) and an F1 score of 0.975 (95% CI: [0.9495, 0.9947]; Wilcoxon test, *p* < 0.001) across the validation dataset. Post-deployment evaluations also showed strong performance of the deployed GPT-4o model with a sensitivity of 1.0 (95% CI: [1.0, 1.0]), a specificity of 0.94 (95% CI: [0.8913, 0.9804]), and an F1 score of 0.97 (95% CI: [0.9479, 0.9908]). This high level of accuracy supports a reduction in manual review, streamlining clinical workflows and improving diagnostic precision.

**Conclusions:** The GPT-4o model provides an effective solution for the automatic extraction of PE diagnoses from radiology reports, offering a reliable tool that aids timely and accurate clinical decision-making. This approach has the potential to significantly improve patient outcomes by expediting diagnosis and treatment pathways for critical conditions like PE.

## Introduction

Pulmonary embolism (PE) is a serious medical condition where a blood clot blocks one of the pulmonary arteries in the lungs, typically originating from a vein in the lower limbs [1–3]. This blockage can significantly impede blood flow, leading to reduced oxygen levels in the blood and potential lung tissue damage. PE is critical because it can cause sudden, life-threatening complications such as cardiac dysfunction and other acute admissions [4,5]. Prompt diagnosis and treatment are crucial to improve outcomes and reduce the risk of mortality [6,7].

Clinical imaging techniques commonly used for diagnosing pulmonary embolisms include pulmonary computed tomography angiography (CTA), combined CT venography and pulmonary angiography (CVPA), and multi-detector CT angiography (MDCTA) [8–10]. The analysis and outcomes of these modalities are recorded in radiology reports which describe the presence or absence of emboli, their location, size, and impact on pulmonary circulation. Radiology reports are structured documents that capture the conditions observed from radiology images [11,12]. Typically, the most important parts of these reports are the Findings and Impression sections [13]. The Impression section provides a clinically precise summary of the patient’s status, typically summarizing the key findings and diagnoses from the Findings section [14]. Therefore, the diagnosis of PE is highly likely to be mentioned in the Impression section. Early documentation of PE and its extraction in the (electronic medical record) EMR system, and consequently in clinical workflows, is crucial for improving patient outcomes. In this study, we aim to develop an advanced transformer-based text classification model to extract PE diagnoses from the Impression section of radiology reports, expediting structured data availability and enhancing the quality of care through evidence-based practices.

Natural Language Processing (NLP) techniques have been increasingly utilized in the field of radiology, particularly in extracting critical information from radiology reports such as diagnoses [15]. Studies have shown that NLP, combined with machine learning and deep learning algorithms, can effectively extract relevant information from radiology reports [16–18]. These techniques enable the automatic identification and extraction of critical findings such as pleural effusion, pulmonary infiltrate, and pneumonia, aiding in the classification of reports consistent with bacterial pneumonia [19]. Furthermore, NLP algorithms have been developed to detect specific findings like acute pulmonary embolism in radiology reports, showcasing the potential of NLP in enhancing diagnostic processes [20,21].

The application of NLP in radiology reports extends to various medical conditions, including pulmonary embolism. Studies have demonstrated the effectiveness of NLP in structuring the content of radiology reports, thereby increasing their value and aiding in the classification of pulmonary oncology according to the TNM classification system, a standard for staging cancer [22]. Additionally, NLP has been used to identify ureteric stones in radiology reports and to build cohorts for epidemiological studies, showcasing the versatility of NLP in medical research [23].

Recent studies have demonstrated the effectiveness of Clinical-Longformer in various clinical NLP tasks. For instance, it has been utilized to identify incarceration status from medical records, showcasing good sensitivity and specificity compared to traditional keyword-based methods [24]. Additionally, Clinical-Longformer has been successfully applied in the classification of clinical notes for automated ICD coding, where it outperformed other models in accuracy [25,26]. This capability to accurately interpret and classify clinical text is crucial for improving healthcare delivery and ensuring proper coding for reimbursement purposes.

On the other hand, advanced versions of the GPT family like GPT-4 and GPT-4o, generative language model, have been recognized for their versatility in clinical applications, particularly in generating, summarizing, classifying, and extracting clinical information [27–33]. Its multimodal capabilities allow it to process not only text but also images and audio, enhancing its utility in diverse clinical settings [34]. GPT-4 has been employed in clinical trial matching, where it automates eligibility screening, thus streamlining the recruitment process for clinical studies [35].

This study aims to develop an LLM-based extraction system to automatically extract pulmonary embolism diagnoses from radiology report impressions. The key contributions of this study are:

- Enhance and accelerate clinical data availability to improve the quality of care through evidence-based approaches.
- Develop a reliable diagnoses extraction system, which examines two technologies: Clinical Longformer and GPT-4o.
- Deploy the developed system as a cloud-based web application, addressing a gap often found in Clinical AI research.
- Evaluate the model both before and after deployment.

## Methods

In this section, we provide a comprehensive overview of the study’s methodology. Subsequently, we explore the text classification approach, followed by a detailed description of the dataset utilized. We then describe the models applied in this research. Additionally, we discuss the deployment pipeline of the selected model. Finally, we outline the evaluation metrics employed to assess the model’s performance.

### Overview

The primary objective of this study is to develop and deploy an AI solution capable of extracting PE diagnoses from radiology report impressions. After defining this goal, the research proceeds through four distinct phases. In the first step, for collecting training and validation datasets, we determined the appropriate data tables in the Virtua Health clinical database and extracted radiology impressions data. This is followed by preprocessing and transforming the data to make it suitable for model development. The second step involves creating and testing two models (Clinical Longformer and GPT-4o), then choosing the one with the best results to proceed. The selected model is then implemented during the third step. In the final step, the performance of the model was tracked in real-world conditions and evaluated on how it affected operational outcomes.

### Radiology Impressions Text Classification

Text classification is a fundamental task in NLP that involves categorizing text into predefined labels based on its content. This task is widely used in applications such as sentiment analysis, spam detection, and medical report classification. Text classification models typically preprocess the data by tokenizing the text and transforming it into numerical representations suitable for machine learning. An NLP approach is then employed to make predictions based on patterns in the text.

Figure 1 illustrates the process of classifying radiology report impressions to identify PE cases, which is adopted in this study. The workflow begins with the extraction of radiology impressions from a clinical database. The impressions are preprocessed by consolidating line-wise text, removing unnecessary spaces, and applying labels to prepare the data for analysis. Following this, two different text classification models are employed: Approach 1 utilizes a Clinical Longformer Model, while Approach 2 involves GPT-4o, a large language model. Both models classify the impressions into two categories: PE and Non-PE. The goal is to determine whether a diagnosis of PE is present in each radiology report impression.

**Figure 1.**
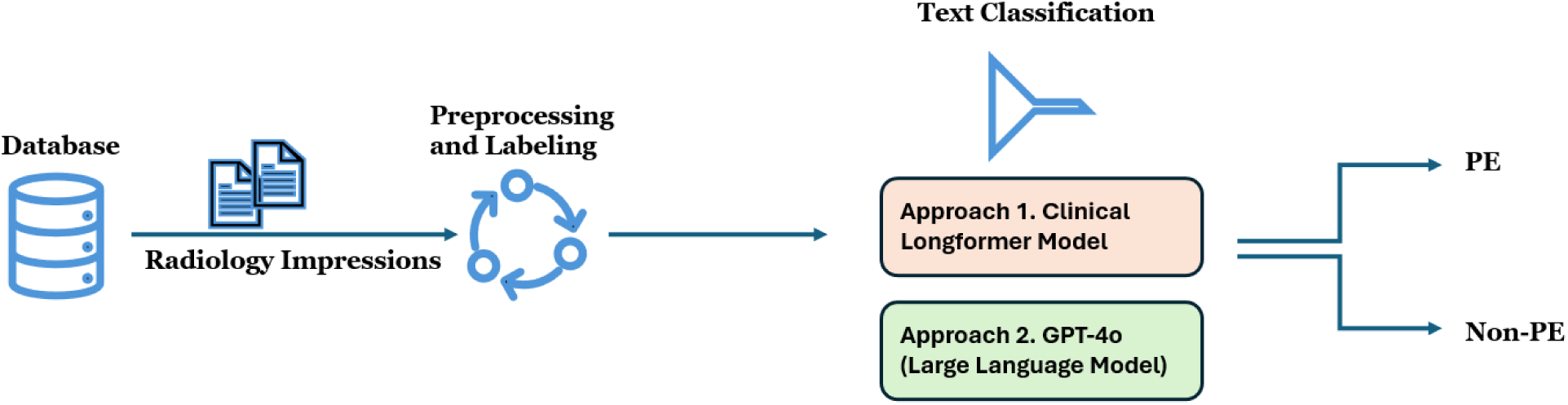
Radiology impressions text classification.

### Data

The data used in this study was sourced from the electronic medical record relational database of Virtua Health, NJ, with the primary data element being the impressions of radiology reports. These impressions, which contain key diagnostic information, were consolidated from line-wise data and cleaned to remove extraneous spaces. This process ensured that the data was formatted appropriately for analysis and modeling.

The training dataset consists of 1,000 samples, which were randomly selected from radiology reports generated between January 1, 2024, and June 30, 2024. For the validation dataset, 200 samples were randomly drawn from radiology reports collected in July 2024. Additionally, a separate testing dataset, consisting of 200 observations, was sampled randomly from operational data received between August 1, 2024, and August 31, 2024. The characteristics of the training and validation datasets are outlined in Table 1. The testing dataset characteristics will be discussed in the following section.

**Table 1.** Training and Validation Data Characteristics.

| **Metric** | **Training Dataset** | **Validation Dataset** |
| --- | --- | --- |
| Number of observations | 1,000 | 200 |
| Average number of words (a token is approximately $\frac{3}{4}$ of a word) | 43.64 | 40.18 |
| Number of pulmonary embolism-positive cases | 36 | 100 |
| Number of pulmonary embolism-negative cases | 964 | 100 |

As shown in Table 1, the training dataset contains 1,000 observations, with an average of 43.64 words (or 32.73 tokens, where a token is approximately three-fourths of a word) per report impression. The training data includes 235 occurrences of pulmonary embolism term, with 36 positive cases for pulmonary embolism and 964 negative cases. The validation dataset, consisting of 200 observations (1:1 ratio of classes), has a slightly lower average word count per report impression, at 40.18 words.

### Fine-Tuned Clinical Longformer Classifier

The Clinical Longformer is a specialized transformer model designed to handle long clinical documents, overcoming the typical limitations of standard transformer models such as BERT, which can process sequences up to 512 tokens [36]. Clinical Longformer incorporates a sparse attention mechanism that allows it to efficiently process sequences up to 4,096 tokens, making it ideal for handling lengthy clinical narratives. Pre-trained on large clinical datasets, it is particularly effective in capturing long-term dependencies in medical text. In this study, the Clinical Longformer is fine-tuned to classify radiology impressions for identifying pulmonary embolism, leveraging its ability to process comprehensive radiology report impressions without truncating important contextual information.

We fine-tuned (full fine-tuning) the Clinical Longformer model on a GPU server with 48 GB of memory. The fine-tuning parameters were as follows:

- Batch size: 4
- Gradient accumulation steps: 8
- Learning rate: 2e-5
- Number of epochs: 5
- Optimizer: AdamW
- Learning rate scheduler: Linear

### GPT-4o Classifier

The methodology for utilizing GPT-4o in the text classification of radiology impressions, specifically for PE diagnosis, is based on a combination of chain-of-thought (COT) reasoning and few-shot learning techniques. As outlined in Figure 2(a), the process begins by initializing an empty list to store the generated labels. GPT-4o is then prompted using a COT and few-shot learning template, where relevant examples of radiology impressions with their corresponding labels (PE or Non-PE) are presented to the model. The temperature parameter is set to zero to minimize randomness in the model’s predictions. For each radiology impression in the dataset, the system inserts the impression into the prompt, calls the GPT-4o API, and receives a response that indicates whether a PE diagnosis is present. The resulting labels are appended to the list for further analysis and validation.

**Figure 2.**
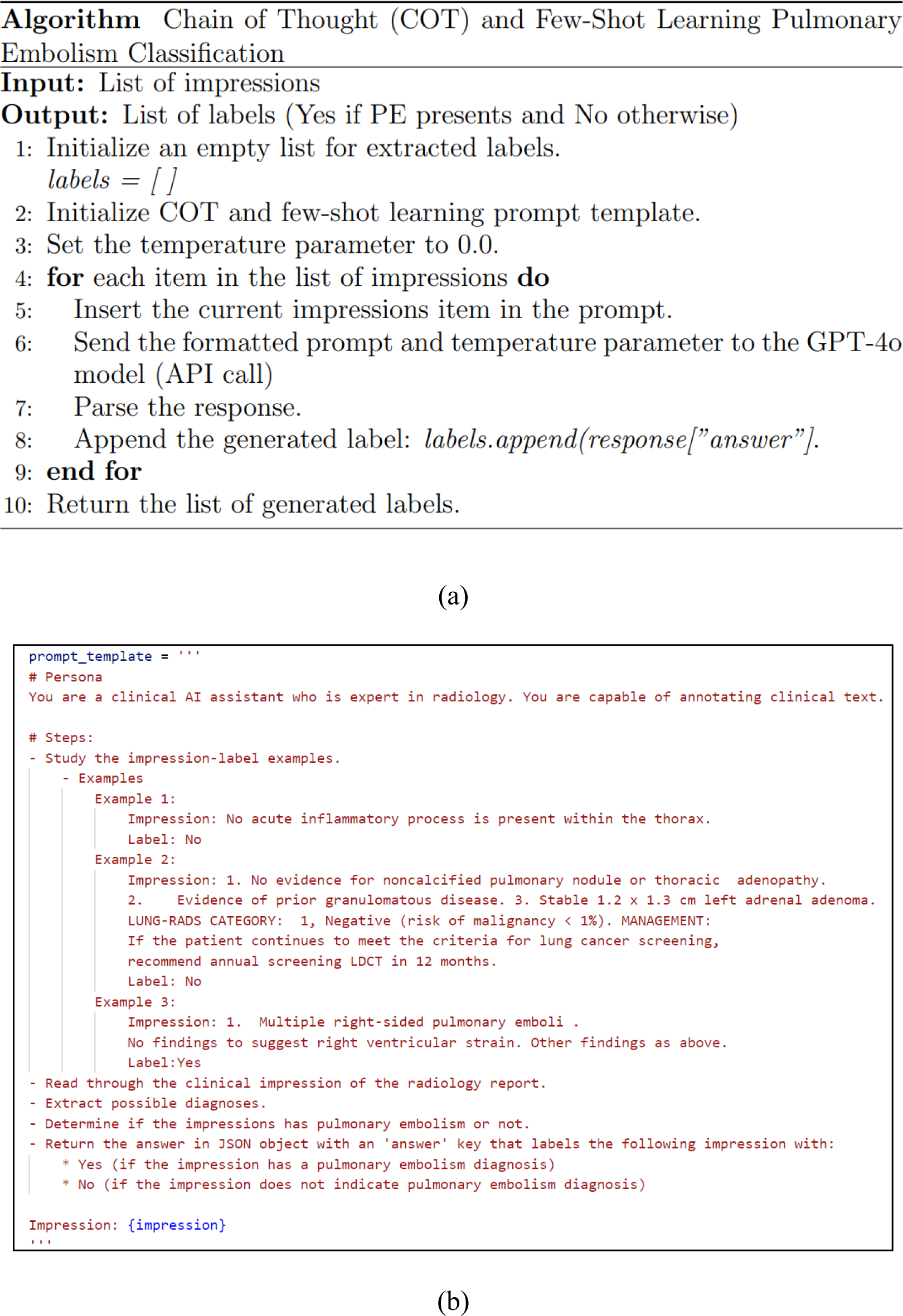
GPT-4o for radiology impressions classification. Extraction of pulmonary embolism diagnosis.

As shown in Figure 2(b), the prompt includes a persona where GPT-4o is defined as a clinical AI assistant proficient in radiology, capable of interpreting complex medical language. The prompt further provides detailed steps, starting with studying example impression-label pairs, followed by reading through the target impression to extract potential diagnoses. The model is tasked with determining whether PE is indicated in the impression and returns the output as a structured JSON object. This methodology leverages GPT-4o’s advanced language comprehension capabilities to classify radiology reports efficiently, using both clinical reasoning and context learned from the few-shot examples.

### Deployment Pipeline

The deployment pipeline for the pulmonary embolism (PE) classification model, illustrated in Figure 3(a), integrates a combination of on-premises and Azure cloud services to create a streamlined and scalable system. The process begins with data being sourced from an on-premises SQL server, which stores radiology report impressions. These impressions are transferred to an Azure SQL database, where they are stored and prepared for further analysis. This architecture utilizes a direct interaction between Azure SQL, an Azure Web App, and the Azure OpenAI service. The Azure OpenAI service, hosting the GPT-4o model, is invoked by the Azure Web App to perform text classification on the radiology impressions and return pulmonary embolism classification results. These results are then stored back in the Azure SQL database. The web app fetches the results from Azure SQL and displays them for end users.

**Figure 3.**
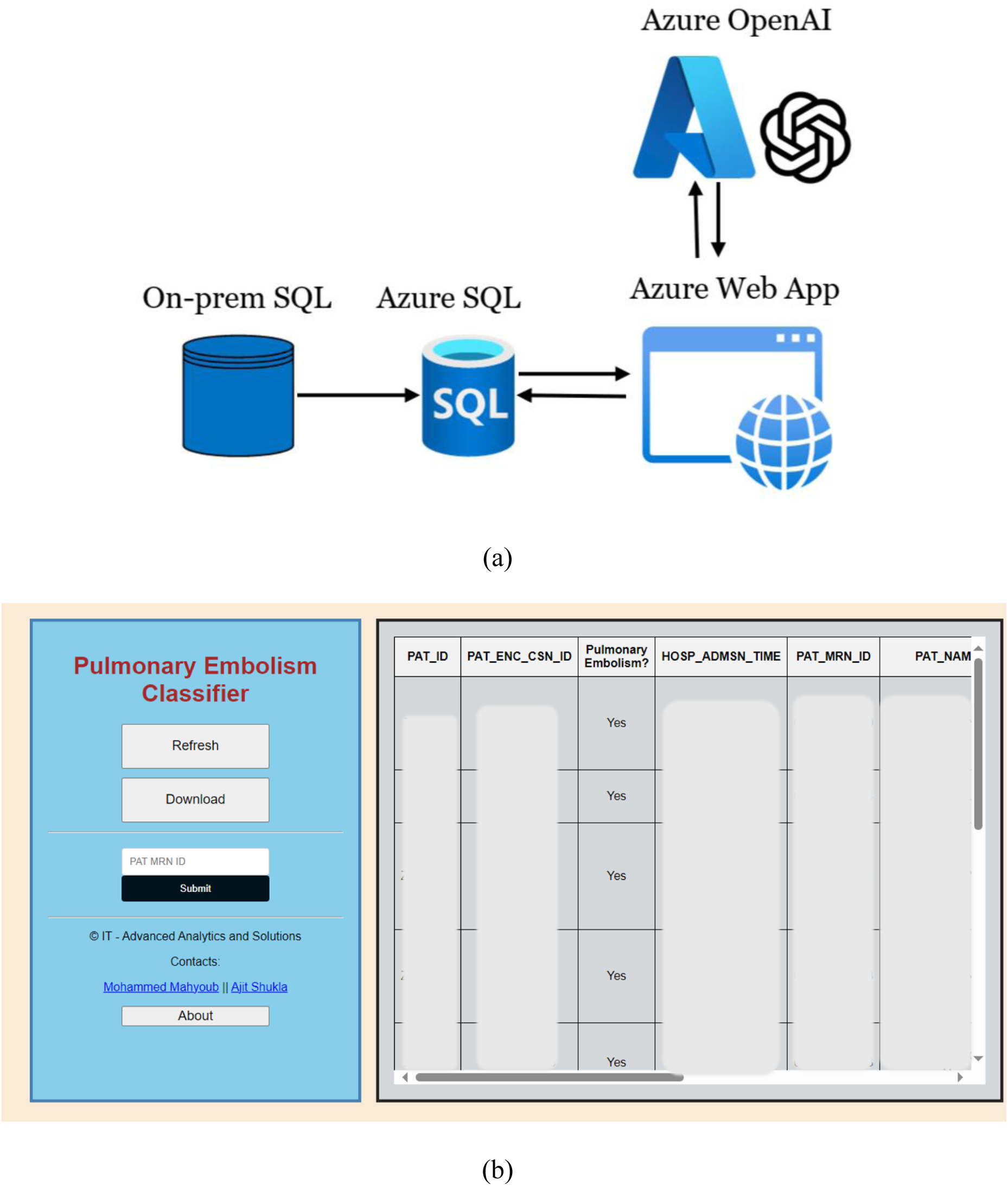
Deployment pipeline and consumption Web App. Only positive cases are displayed. This Web App has been operationalized at Virtua Health, NJ.

As shown in Figure 3(b), the web app was built using Python Flask for the backend, along with HTML, CSS, and JavaScript for the frontend. The interface allows users to query the system by submitting a patient’s medical record number to retrieve the corresponding PE classification result. Users can also refresh the data or download the results for further analysis. The table on the right displays relevant patient information, including patient IDs, encounter IDs, admission times, and the PE classification results. This interface serves as a convenient tool for healthcare professionals to quickly identify patients with a PE diagnosis, improving clinical decision-making and patient outcomes by providing prompt, automated insights. The Web App has been operationalized at Virtua Health, NJ.

### Evaluation Metrics and Statistical Testing

To evaluate the performance of the PE classification model, we employed several commonly used metrics:

- *Sensitivity (Recall)*: The proportion of actual PE cases that the model correctly identified. It measures the model’s ability to detect positive cases (PE) and is defined as the ratio of true positives to the sum of true positives and false negatives.
- *Specificity*: The proportion of actual non-PE cases that the model correctly identified. It reflects the model’s ability to avoid false positives, calculated as the ratio of true negatives to the sum of true negatives and false positives.
- *F1 Score*: A harmonic mean of precision and recall, which provides a balanced measure of the model’s performance, especially in cases of imbalanced data. It is particularly useful for evaluating the trade-off between precision and recall in the context of PE classification.

To rigorously evaluate and compare model performance, we used non-parametric bootstrap sampling with 1,000 iterations on both the validation and post-deployment datasets. In each bootstrap iteration, samples were drawn with replacements from the dataset, and standard evaluation metrics—sensitivity, specificity, and F1 score—were calculated. This process yielded 1,000 metric estimates per model (GPT-4o and Clinical Longformer), providing an empirical distribution for each metric.

From these distributions, the mean and 95% confidence intervals were derived using percentile-based estimation. This approach enables robust quantification of performance uncertainty without assuming normality.

To statistically compare the models, we conducted paired, two-sided Wilcoxon signed-rank tests on the bootstrapped metric distributions. This non-parametric test assesses whether the differences in paired metric values across bootstraps are statistically significant. A significance level of α = 0.01 was used to determine the threshold for significance.

The same bootstrapping procedure was applied to the post-deployment dataset to calculate mean metric values and corresponding 95% confidence intervals. However, statistical significance testing was conducted only on the validation dataset to ensure controlled model comparisons under consistent evaluation conditions. In the post-deployment setting, only the GPT-4o model was in use.

## Results

This section presents the research findings. First, we evaluate the Clinical Longformer and GPT-4o models using the validation dataset during the development phase. Then, we assess the performance of the deployed GPT-4o model post-deployment.

### Models Evaluation

Figure 4 presents a comparative analysis of evaluation metrics, including sensitivity, specificity, and F1 score, between GPT-4o and Clinical Longformer. The results demonstrate statistically significant differences across all three metrics (*p*<0.001), as determined by the Wilcoxon signed-rank test.

**Figure 4.**
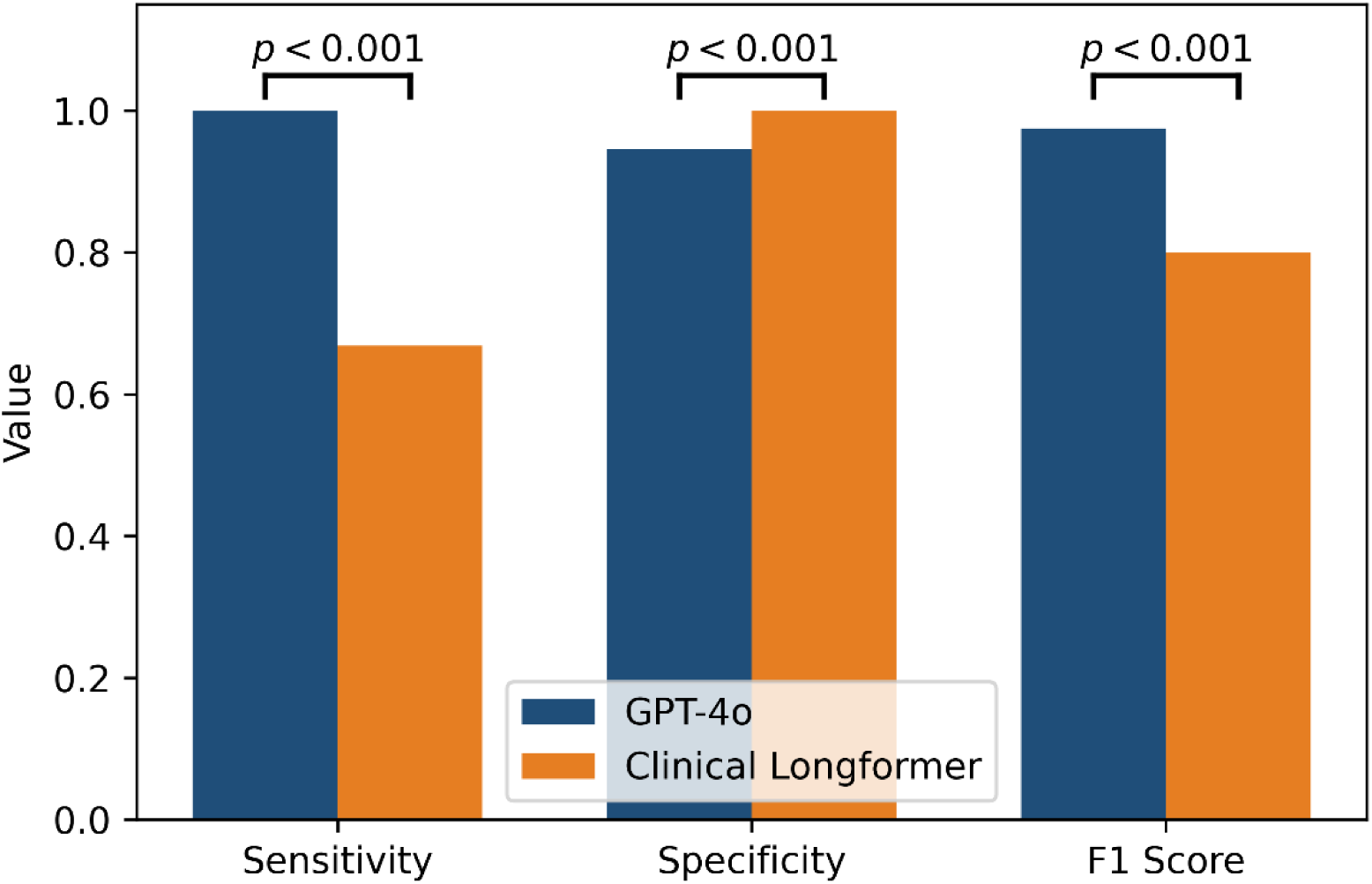
Evaluation metrics comparison of Clinical Longformer (baseline model) and GPT-4o.

GPT-4o exhibits superior sensitivity, achieving a perfect sensitivity of 1.0 (95% CI: 1.0–1.0), in contrast to Clinical Longformer, which attains a sensitivity of 0.6684 (95% CI: 0.5644–0.7604). This indicates that GPT-4o captures all positive cases correctly, whereas Clinical Longformer has a notable false negative rate. For specificity, Clinical Longformer achieves a perfect score of 1.0 (95% CI: 1.0–1.0), slightly surpassing GPT-4o, which attains a specificity of 0.9456 (95% CI: 0.9022–0.989). Although both models perform well in distinguishing negative cases, GPT-4o exhibits a marginally higher false positive rate. Regarding the F1 score, GPT-4o significantly outperforms Clinical Longformer, with an F1 score of 0.975 (95% CI: 0.9495–0.9947) compared to 0.8002 (95% CI: 0.7215–0.8639) for Clinical Longformer. This improvement reflects GPT-4o’s higher balance between precision and recall, leading to superior overall classification performance.

The statistically significant differences across all three metrics underscore GPT-4o’s robust generalization and enhanced performance in comparison to Clinical Longformer, suggesting its potential for improved clinical applications.

### Post-Deployment Performance

Based on GPT-4o’s excellent performance during the validation phase, where it achieved superior metrics, it was selected for deployment in the operational setting. The post-deployment evaluation of the GPT-4o model was conducted using a dataset of 200 records, which was selected using stratified sampling from the operational data. Table 2 provides a summary of the dataset characteristics, including 100 positive cases of pulmonary embolism and 100 negative cases.

**Table 2.**
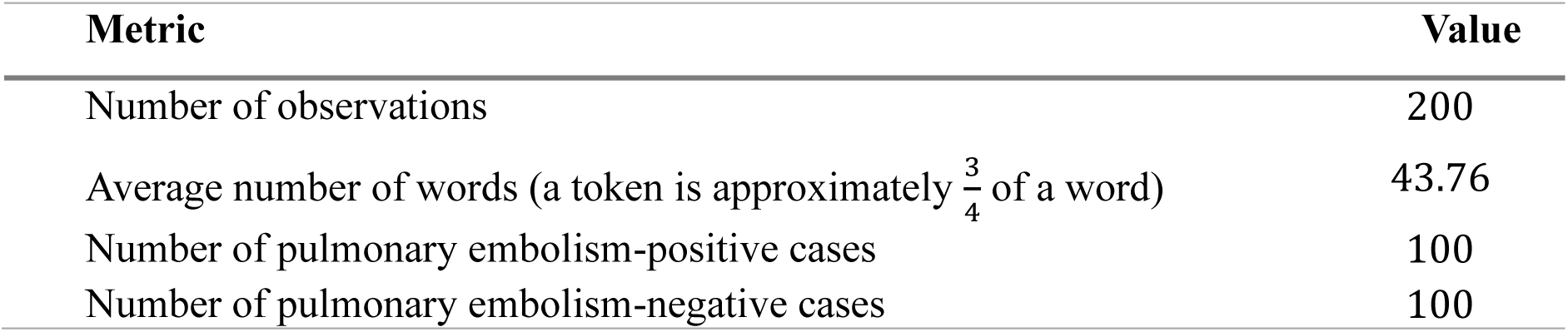
Post-Deployment Testing Data Characteristics. A stratified random sample of 200 records was taken from the operational dataset for post-deployment evaluation.

Figure 5 presents the post-deployment performance metrics of GPT-4o, including sensitivity, specificity, and F1 score, along with their respective 95% confidence intervals. Sensitivity remains at a perfect 1.0 (95% CI: [1.0, 1.0]), demonstrating the model’s ability to correctly identify all positive cases without false negatives. Specificity, however, shows a slight decrease compared to pre-deployment values, with a point estimate of approximately 0.94 (95% CI: [0.8913, 0.9804]), suggesting a modest increase in false positives. The F1 score remains high at approximately 0.97 (95% CI: [0.9479, 0.9908]), indicating a strong balance between precision and recall. While GPT-4o maintains robust predictive performance post-deployment, with high sensitivity and a stable F1 score, the slight decline in specificity highlights a potential area for further refinement.

**Figure 5.**
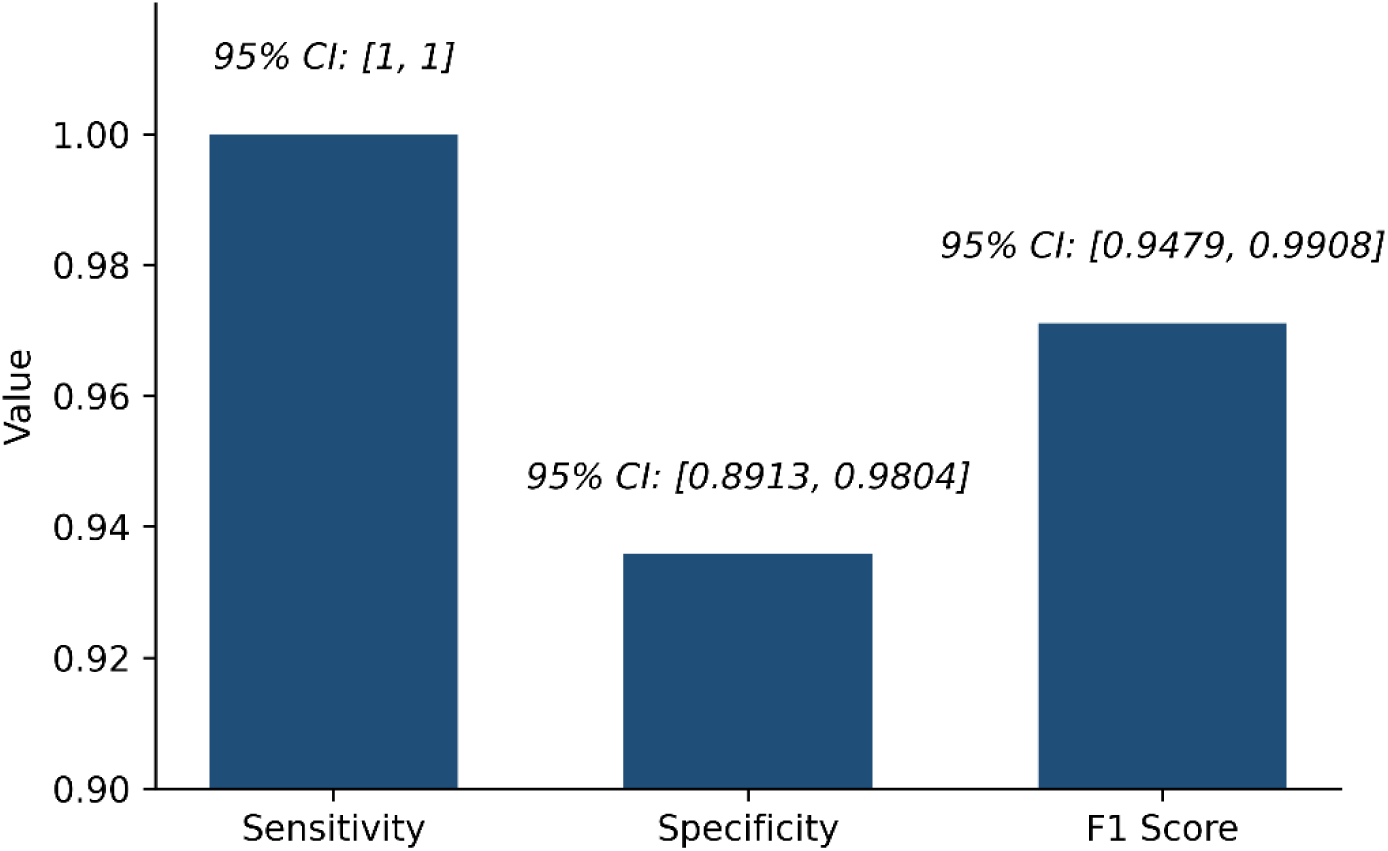
Post-deployment performance metrics of GPT-4o.

## Discussion

### Principal Findings

This study underscores the efficacy of GPT-4o in automating the extraction of PE diagnoses from radiology report impressions. GPT-4o demonstrated excellent performance across sensitivity, specificity, and F1 scores during both validation and post-deployment evaluations. These findings suggest that GPT-4o’s advanced language understanding capabilities allow it to capture subtle contextual and semantic nuances in radiology impressions, which are often critical for accurate diagnosis.

This success is attributed to GPT-4o’s ability to leverage large-scale training on diverse datasets, which enhances its generalizability and adaptability. Additionally, its deployment as a cloud-based tool offers scalability, making it accessible to healthcare systems of varying sizes. These attributes make GPT-4o a transformative tool in clinical AI, setting a benchmark for future applications in radiology and other diagnostic domains. This success has also been recognized in various studies, as GPT-4o has demonstrated promise in addressing critical tasks across different areas of radiology [37–41].

### Error Analysis

As indicated in the results section, GPT-4o scored 1.0 (95% CI: [1.0, 1.0]) on sensitivity. However, the specificity was 0.94 (95% CI: [0.8913, 0.9804]), hinting at a false positive rate of around 6%. Here, we look at three of these false positives to uncover any commonality.

The first sample, labeled as “Yes” while the ground truth (human labeler) says otherwise, has the following impressions text: “Mild right upper lobe infiltrate. Heterogeneous density of the pulmonary artery, which could represent pulmonary embolus.” This is a borderline case, where the text indicates a likelihood of PE without firm confirmation. The model was able to bring attention to this case for further evaluation. Also, ground truth labeling may suggest the need for further validation.

The second sample’s impressions text states “1. Limited examination as mentioned above, particularly further evaluation of the pulmonary arteries in the right lung. Questionable subsegmental pulmonary embolism may be present in the right middle lobe. Follow-up would be helpful as clinically indicated. 2. Large right pleural effusion. Adjacent lung consolidation suggesting atelectasis. 3. Mild groundglass infiltrate in the right upper lobe and left lower lobe. This may represent mild pneumonitis. 4. Small left pleural effusion. 5. Incidental findings as above.” The model indicated the presence of PE. If we look at the text, we can see that the radiologist indicated a questionable PE. This case is similar to the previous one, in which the model labels likelihoods and questionable diagnoses as indicative of PE, which could be a useful way to bring clinicians’ attention to the case for further evaluation.

The third sample impressions text is “1. Small linear filling defects involving the right lower lobe, left upper lobe, and left lower lobe, likely sequela of chronic pulmonary embolism. No convincing findings of acute pulmonary embolism. No imaging findings of right heart strain. 2. Interval enlargement of multiple pulmonary nodules measuring up to approximately 12 mm. Further correlation with PET/CT versus follow-up CT in 3 months is recommended. 3. Areas of mild subpleural atelectasis or scar with mild interstitial thickening that may be related to chronic change, although mild pulmonary edema or infection would be difficult to entirely exclude.” The text indicates that “No convincing findings of acute pulmonary embolism.” This resulted in a negative ground truth label. However, the model interpreted the “likely sequela of chronic pulmonary embolism” as an indicator of PE. The latter suggests that filling defects were a result of a past medical condition. Thus, this might be a definite false positive.

In summary, analyzing some of the false positives indicates the model’s ability to bring attention to borderline cases that might hint at the presence of PE, allowing for further clinical validation. However, the model sometimes results in definite false positives when the radiology text ambiguously discusses PE in the context of past conditions.

### Operational and Clinical Implications

The post-deployment results of GPT-4o show several important operational and clinical implications. First, the model’s sensitivity and specificity in a real-world setting indicate that it can help distinguish between subjects who have pulmonary embolisms and those who do not, without many false positives or negatives. This is important in a clinical setting where missing cases of pulmonary embolism have implications for patient outcomes, and false positives can lead to unnecessary further investigation or treatment. At Virtua Health, the model successfully identified over 700 positive PE cases in 2024.

From an operational perspective, the model’s excellent accuracy on both the validation and post-deployment testing datasets means that the model can be operated in an autonomous manner which would help reduce the workload of radiologists and other healthcare professionals. As a result, it can classify cases correctly, thus taking some of the pressure and time off clinicians and their ability to concentrate on other, more challenging or time-consuming cases. Furthermore, the model’s high precision means that healthcare resources are used more effectively, with fewer unnecessary interventions and therefore better patient care.

Clinically, the use of GPT-4o increases the decision support that clinicians can receive by offering a correct answer to the question regarding the presence of pulmonary embolism based on the information presented in the radiology reports. This early recognition can result in early diagnosis and management and therefore better patient results and possibly reduced mortality. Moreover, the stability of the performance of GPT-4o in identifying pulmonary embolisms in various datasets suggests robustness and generality, which could be useful in various clinical settings.

### Ethical Considerations

The employment of AI models in healthcare induces multifaceted ethical issues that need to be deliberated on. The first and foremost concern is the privacy of the patients and the security of the information because medical information is private. This includes strong network security, policy compliance with laws like HIPAA and GDPR, and data exposure minimization during training and use of the model. It is crucial to ensure that AI systems work within these legal and ethical parameters to ensure the credibility of their use.

Another important factor is compliance with regulations because the application of AI in healthcare has to be compatible with the standards that are set on a regional and international level. This includes making sure that the process of training, validation, and integration into the clinical workflow of the model is well documented. These processes are easily understandable and can be explained to healthcare providers, patients, and regulators, thus establishing the ethical and legal sustainability of technology.

Another important issue is bias and fairness as AI models trained on small and biased datasets are likely to emit biased output. For instance, if the training data is biased then the predictions of the model may be unfair to some patients and thus lead to unequal care. This is particularly a concern in healthcare where discrimination can have severe impacts on minority populations, stressing the need to ensure that datasets are inclusive, and model evaluation focuses on fairness.

Finally, it is important that in using AI in the various fields of life, transparency and accountability are considered to create confidence in the use of AI. The results of the model must be easy to understand and explain to the clinicians and patients, hence the need for explainable AI. Moreover, it is crucial to subject the models to routine assessments and performance tracking to provide evidence of accountability and trustworthiness in the long run.

To address these ethical issues, there is a need for ongoing assessment, open reporting, and interaction between AI creators, healthcare deliverers, and regulators. Thus, the main aspects that can be improved are the aspects that when improved will help achieve the best possible results with the use of AI in healthcare while at the same time adhering to ethical principles and protecting the rights of patients.

### Bias Evaluation

LLMs are generally pretrained on large corpora of texts from the internet. As a result, inherent biases in human language can influence model outputs. Recent LLMs, such as GPT-4o, undergo post-training alignment to reduce biased responses. This serves as the first line of defense against bias. Secondly, in our case, the LLM is instructed to return a single label (Yes or No), which may further reduce the potential for hallucinations and bias. LLMs typically require more tokens to reveal their underlying tendencies, which can include bias from the pretraining stage. Lastly, we evaluated the model’s performance using the F1 score across different age groups and gender profiles.

Table 3 summarizes the F1 score across different sex and age groups to assess the consistency of the model’s performance across demographic subpopulations. The model demonstrates comparable F1 scores for both female (0.970, 95% CI = [0.930, 1.000]) and male (0.972, 95% CI = [0.937, 1.000]) groups, indicating no apparent sex-based disparity in classification performance.

**Table 3.** F1 score across sex and age groups.

| Variable | Group | Count (Percentage) | F1 Score |
| --- | --- | --- | --- |
| Sex | Female | 98 (51%) | 0.970 (95% CI = [0.930, 1.000]) |
|  | Male | 102 (49%) | 0.972 (95% CI = [0.937, 1.000]) |
| Age | <30 | 10 (5%) | 1 (95% CI = [1, 1]) |
|  | 30-50 | 25 (12.5%) | 0.963 (95% CI = [0.867, 1.000]) |
|  | 50-70 | 93 (46.5%) | 0.982 (95% CI = [0.951, 1.000]) |
|  | 70-90 | 62 (31%) | 0.966 (95% CI = [0.906, 1.000]) |
|  | 90+ | 10 (5%) | 0.823 (95% CI = [0.500, 1.000]) |

Across age groups, the F1 scores remain generally high, with minor variations. The highest score is observed in individuals under 30 years old (F1 = 1.000), although this group comprises only 5% of the sample, suggesting potentially limited generalizability due to the small sample size. The model performs well for the majority age groups—50–70 years (F1 = 0.982, 95% CI = [0.951, 1.000]) and 70–90 years (F1 = 0.966, 95% CI = [0.906, 1.000])—which collectively represent over 75% of the dataset. However, performance decreases slightly for individuals aged 90 and above (F1 = 0.823, 95% CI = [0.500, 1.000]), likely due to a smaller sample size and increased clinical complexity in this cohort. Overall, the model demonstrates robust and consistent performance across most demographic groups, with slight variation observed at the extremes of age distribution.

### Explainability

GPT-4o is an LLM built on the transformer architecture, a type of neural network that uses an attention mechanism to learn semantics. Due to its complexity, the model is inherently a black box. However, since the model generates output by continuing the input text based on user instructions, the output can be interpreted as the most probable completion (especially when the temperature parameter is set to 0.0).

In our case, the instructions are to analyze radiology impressions and return a Yes or No, indicating the presence or absence of PE, respectively. In real-world decision-making, users are informed that the LLM generates responses based on the most likely continuation of the input text according to the instructions provided. The generated label is returned to users along with the corresponding impressions for validation and confirmation.

Exploring explainable AI (XAI) methods is highly encouraged to better understand how input text influences LLM output, improving predictability and transparency. Techniques such as Shapley Additive Explanations (SHAP) and attention mechanism analysis can support this effort. In our case, implementation was challenging due to the use of a large proprietary LLM (i.e., GPT-4o), for which we do not have access to the model architecture needed for such analyses. Therefore, leveraging open-source LLMs in the future could enable the application of these techniques.

The GPT-4o-based PE classifier is delivered through a web application, requiring user involvement to confirm the model’s results. In summary, our application adopts a human-in-the-loop approach. While this supports practical use, the broader explainability of LLM outputs remains an ongoing area of research aimed at better understanding the underlying mechanisms of next-token prediction.

### Limitations

As the study shows promising results, there are important limitations, and their implications cannot be overlooked. A major limitation is the size of the dataset used in this study which is 1,000 training samples, 200 validation samples, and 200 post-deployment testing samples. Another limitation is the data imbalance in the training dataset (for the Clinical Longformer). With such a restricted number of examples, the model may not be able to generalize its learning to the full extent of the clinical scenarios that are likely to be encountered in the real world. This could lead to it being less accurate in real-world use, where report structures can differ significantly, and larger datasets are usually used to increase model confidence and stability.

Another limitation has to do with the model’s generalization. The training data was collected from a single healthcare organization, Virtua Health, which means that the language and cultural features that are unlikely to be encountered in reports from other hospitals or countries may not be well-represented in the model. As a result, this may restrict the applicability of the model to other settings unless it is retrained (or prompt-engineered) with local data.

The study also found that the Clinical Longformer, which was employed as a baseline, could not grasp some of the semantic details, resulting in the misclassification of two positive pulmonary embolism cases. This suggests that future work should build on the current study to enhance the architecture of transformer models specific to clinical tasks, especially those that entail processing medical text with sophistication.

Some issues may occur when implementing the model in the real world even though GPT-4o performed excellently in controlled experiments. Real-world data often contains errors, omissions, or other noise that the model may not be able to handle properly. In addition, there are potential threats to consistency in reporting practices that may lead to inconsistencies in the information captured by the model.

### Future Work

To overcome these limitations, future work should focus on the growth of the dataset, which should be expanded to include a bigger and more complex set of samples that would encompass different types of radiology reports, patients, and institutions. This will increase the size of the dataset and the variety of the samples, which in turn will increase the model’s coverage and stability. Data imbalance in the training dataset of the Clinical Longformer should be tackled by using data augmentation techniques to test the impact of validation performance. Furthermore, the model must be tested in different healthcare contexts to determine its effectiveness, which means that external validation is critical. External validation can be achieved through collaborations with other health systems or testing on publicly available datasets. To mitigate real-world operationalization challenges, future implementation should include routine performance audits to monitor model accuracy over time, periodic retraining with updated real-world data to maintain robustness, and regular human validation checks to ensure data quality and reflect evolving documentation practices. These strategies can help maintain reliability despite variability in data quality and reporting standards. Overall, these measures will assist in ensuring that the model is not only efficient but also easily implementable in various healthcare environments.

### Conclusions

In conclusion, this study features an effective LLM-based approach to the automation of the extraction of pulmonary embolism (PE) diagnoses from radiology report impressions. We then compared the performance of a fine-tuned Clinical Longformer and GPT-4o and found that GPT-4o outperformed in terms of sensitivity, specificity, and overall accuracy both pre and post-deployment. The integration of GPT-4o into clinical practice has several operational and clinical benefits, including decreasing the need for manual review, improving clinical decision support, and detecting cases of PE more quickly. Furthermore, the model’s capability to lighten the load of manual review to a great extent is a key contributor to improving the workflow of the diagnostic process, thereby allowing clinicians to channel their efforts toward more challenging problems. These qualities make it suitable for application in other clinical settings with the potential to go beyond PE diagnosis to other medical conditions that require comprehensive and accurate analysis of radiology reports. Not only does its integration enhance efficiency but also the quality of clinical decision-making processes. Future work may involve expanding this approach to other medical conditions and improving the integration of NLP-based models into clinical workflows to keep on enhancing the quality of healthcare delivery.

## Author Contributions

*Mohammed Mahyoub*: Conceptualization, Data curation, Formal analysis, Investigation, Methodology, Software, Validation, Visualization, Writing – original draft, Writing – review & editing. *Kacie Dougherty*: Writing – review & editing, Formal analysis, Validation. *Ajit Shukla*: Project administration, Resources, Writing – review & editing, Validation.

## Data Availability

The data analyzed in this study is subject to the following licenses/ restrictions: data in the present study are not available due to agreements made with the IRB of Virtua Health.

## Ethics Statement

The studies involving humans were approved by Virtua Health Institutional Review Board FWA00002656. The studies were conducted in accordance with the local legislation and institutional requirements. The ethics committee/institutional review board waived the requirement of written informed consent for participation from the participants or the participants’ legal guardians/next of kin because the research involved no more than minimal risk to subjects, could not practically be carried out without the waiver, and the waiver will not adversely affect the rights and welfare of the subjects. This requirement of consent was waived on the condition that, when appropriate, the subjects will be provided with additional pertinent information about participation.

